# The neutrophil-to-lymphocyte ratio, as an emerging marker, is an important indicator predicting spontaneous reperfusion and clinical prognosis in patients with ST-segment elevation myocardial infarction

**DOI:** 10.1101/2023.03.07.23286964

**Authors:** Bing Li, Mingyou Zhang, Yaoting Zhang, Yang Zheng, He Cai

## Abstract

**Background:** Neutrophil to lymphocyte ratio (NLR) has emerged as a new inflammation marker, which plays a major role in plaque instability, rupture, and erosion, and facilitates its progression, leading to acute myocardial infarction. The study aims to explore the role of NLR in predicting spontaneous reperfusion (SR) and prognosis in patients with ST-segment elevation myocardial infarction (STEMI).

**Methods:** This was a retrospective analysis including 506 STEMI patients undergoing primary percutaneous coronary intervention treatment, who were divided into two groups according to the thrombolysis in myocardial infarction (TIMI) flow: SR group (69 patients, initial TIMI flow 3) and No-SR group (437 patients, initial TIMI flow 0-2).

**Results:** The incidence of SR was 13.6%. SR group was associated with a remarkably lower level of NLR [5.14 (2.97, 7.02) vs. 8.03 (4.54, 10.92), P<0.001], more proportions of final TIMI 3 flow (98.6% vs. 91.5%, P < 0.05), lower incidence of congestive heart failure (8.7% vs. 18.5%, P < 0.05), and significantly better outcomes. Using multivariate logistic regression analysis, NLR (OR: 0.799, 95% CI: 0.730-0.874, P < 0.001) and fasting blood glucose were the independent predictors of SR. On multivariate Cox regression analysis, NLR (HR: 1.035, 95% CI: 1.001-1.071, P < 0.05) was the independent predictor of MACEs during follow-up.

**Conclusions:** NLR had the ability in predicting SR in STEMI patients and SR flow was associated with a favorable outcome. We also revealed an association between NLR and increased risk of MACEs during follow-up.

**Clinical Perspective:** *What Is New?:*

- The incidence of spontaneous reperfusion was 13.6%. Patients with spontaneous reperfusion had a remarkably low level of NLR [5.14 (2.97, 7.02) versus 8.03 (4.54, 10.92), P<0.001], more proportions of final TIMI 3 flow (98.6% versus 91.5%, P < 0.05), lower incidence of congestive heart failure (8.7% versus 18.5%, P < 0.05), and favorable outcomes.
- NLR was not only an independent predictor of spontaneous reperfusion, but also was the independent predictor of major adverse cardiac events during follow-up (HR: 1.035, 95% CI: 1.001-1.071, P < 0.05) in patients with ST-segment elevation myocardial infarction.

*What Are the Clinical Implications?:*

- The level of neutrophil to lymphocyte ratio in patients with ST-segment elevation myocardial infarction is associated with low occurrence of spontaneous reperfusion and adverse outcomes, although, the patients received primary percutaneous coronary intervention.
- Neutrophil to lymphocyte ratio the plays a major role in the risk classification of patients with ST-segment elevation myocardial infarction.

## Introduction

Acute myocardial infarction (AMI) is the most catastrophic presentation of atherosclerotic coronary artery disease, and it’s one of the leading causes of mortality and morbidity worldwide. ^1^ ST-segment elevation myocardial infarction (STEMI), the most acute manifestation of AMI, usually results from a total occlusion of an epicardial coronary artery caused by a thrombus (a blood clot). ^2^

The main therapy for patients with STEMI is early reperfusion to restore epicardial coronary blood flow of the infarct-related artery (IRA). Fortunately, it is appreciated that some patients experience early spontaneous reperfusion (SR) before receiving reperfusion therapy (i.e., intravenous thrombolysis or percutaneous coronary intervention [PCI]). ^3^ The presence of thrombolysis in myocardial infarction (TIMI) 3 flow in the IRA before receiving reperfusion therapy is defined as SR, which is associated with an excellent prognosis. ^4-6^

In the pathogenesis of atherosclerosis, inflammation plays a major role in plaque instability, rupture, and erosion, and facilitates its progression, which subsequently leads to thrombus formation and results in coronary artery occlusion and myocardial infarction. ^7,8^

A great many of inflammatory cells, such as neutrophils, aggregate in the vessel wall, and infiltrate into the atherosclerotic plaques, promoting vascular endothelial cells adhesion and mediating destabilization of atherosclerotic plaques. ^9,10^ Lymphocytes have been reported to play a significant role in mediating the inflammatory response at all phases of the atherosclerotic process and can lead to rupture of atherosclerotic plaques. ^9^

Recently, increased NLR was reported to be associated with malignant tumors, ^11^ inflammatory disease, ^12^ stoke, ^13^ and other diseases . In addition, both high neutrophil counts and lymphocytopenia were found to be associated with cardiovascular disease, ^14,15^ hence, neutrophil to lymphocyte ratio (NLR) has emerged as a new inflammation marker, which can better reflect the state of systematic inflammation.

However, as far as we know, the predictive value of NLR on the SR and long-term major cardiovascular events (MACEs) in STEMI patients has been rarely described. In this study, we aimed to investigate the role of NLR in predicting SR and prognosis in patients undergoing primary PCI due to STEMI.

## Methods

### Study Participants

This study was a single-center, retrospective, and observational study, approved by the Ethics Committee of the First Hospital of Jilin University. In total, 506 patients admitted to the First Hospital of Jilin University from January 2014 to December 2014. The patients were divided into two groups according to the initial TIMI flow: No-SR group (TIMI 0-2 flow on the initial angiogram) and SR group (TIMI 3 flow on the initial angiogram). Inclusion criteria: 1. Age > 18; 2. Diagnosed as STEMI; 3. Underwent primary percutaneous coronary intervention (stent implantation or PTCA) within 12 hours since symptoms onset. Patients with clinical evidence of cancer, autoimmune disease, severe infection, hepatic failure, thrombolytic therapy, hematologic disease, and incomplete clinical data were excluded from the study. To further determine the association of NLR and MACEs, all patients were divided into low NLR group (NLR ≤ 6.57, n=253) and high NLR group (NLR > 6.57, n=253) according to the median of NLR.

### Study variables

Data on gender, age, cardiovascular risk factors (smoking status, hypertension, and diabetes mellitus), physical examination (heart rate, systolic blood pressure, and diastolic blood pressure) and medical history were recorded. Blood samples for baseline laboratory parameters were drawn immediately upon admission, before PCI. Lipid profiles and glucose were also measured in all patients after an overnight fast. Transthoracic echocardiography was routinely performed on each patient within 24 hours of admission to assess cardiac function.

### Angiographic analysis

All participants received a chewable acetylsalicylic acid (loading dosage 300 mg) and clopidogrel (300 mg loading dosage) at admission, and unfractionated heparin 100 U/kg intravenously before the procedure. After angioplasty, all patients were admitted to the coronary care unit where routine antithrombotic therapy was given as a daily dose of 100 mg of aspirin, 75 mg of clopidogrel, subcutaneous administration of enoxaparin. During hospitalization, ACE inhibitor, β-blocker, and statin were initiated in all patients unless contraindicated. Drug-eluting stents were the only stents preferred. Primary angioplasty was performed only for IRA. Patients’ angiographic data were reached by assessing catheter laboratory records. Lesion with ≥ 50% diameter stenosis in vessels ≥1.5 mm in diameter was defined as the diseased vessel. The IRA was classified according to the Thrombolysis In Myocardial Infarction (TIMI) classification ^16^ before and after a procedure.

### Clinical follow-up and outcomes

Clinical follow-up was performed at the first month and every 6 months after index PCI. Data about patients were obtained through out-patient examination and telephone or the statewide death registry database. Primary end points were long-term all-cause mortality, major adverse cardiovascular events (MACEs) including death from any cause, non-fatal re- infarction, repeated ischemic with target vessel revascularization, typical symptoms of angina pectoris, gastrointestinal bleeding, stroke and intracranial hemorrhage during 30-month clinical follow-up.

### Statistical analysis

For categorical variables, data was described as numbers (percentages). The continuous variables with a normal distribution were presented as the mean ± standard deviation, and those with a non-normal distribution were presented as the quartiles [median (quartile 25, 75%)]. Kolmogorov-Smirnov’s test was utilized to evaluate those continuous data’s normality. The statistical difference between continuous data were compared by Student’s t-test for normally distributed values and Mann-Whitney U test for those non- normally distributed. Categorical variables were compared using Fisher’s exact or Chi-square test. Variables associated with SR at *P* < 0.10 in univariate analysis were selected to multivariate logistic regression analysis to identify independent factors strongly associated with SR. To further determine the association of NLR and initial TIMI flow, we performed P for trend and Per SD analysis. NLR was categorized in quartiles: quartile 1, ≤4.23; quartile 2, 4.24-6.67; quartile 3, 4.24-6.67; and quartile 4, ≥10.18. Binary logistic regression analysis was used to estimate the ORs and 95% CIs of type initial TIMI 0-2 flow with NLR quartiles and quantify the linear trend. We also calculated the OR and 95% CI of initial TIMI 0-2 flow associated with per SD increment in the logarithmic transformed NLR. Logistic regression models were performed to estimate the odds ratio (OR) and 95% confidence interval (CI). Receiver operating characteristic (ROC) curve was used for analyzing the cut-off value of NLR. Kaplan-Meier analysis was constructed with the log-rank test to contrast the prognosis during follow-up. To identify independent predictors for cumulative MACEs, variables with *P* < 0.10 in univariable Cox regression analysis were entered into multivariate Cox regression analysis. Hazard ratios and 95% confidence intervals were identified. All of the analyses were 2-tailed. For all tests, *P* < 0.05 was considered statistically significant.

To further demonstrate the association between NLR and SR, the predictors will be showed in a nomogram. The patients were randomly divided into training cohort (n=353) and validation cohort (n=153) by the rate of 7 to 3. NLR was categorized in tertiles: T 1, ≤ 5.15; T 2, 5.16-8.41; T 3 ≥ 8.42. FBG was divided in tertiles: T 1, ≤ 5.71 mmol/L; T 2, 5.72-7.56 mmol/L; T 3 ≥ 7.57 mmol/L. The construction of nomogram was performed by running the programming codes and a series of Stata packages (lrtest, roctab, hl, pmcalplot, dca, nomolog, etc.), which were downloaded and installed from Stata Software online platform. The nomogram’s discriminant performance was assessed and analyzed by receiver operating characteristic (ROC) and area under curve (AUC). The model calibration was assessed by calibration curve and Hosmer-Lemeshow test. Decision curve analysis (DCA) was performed to evaluate the clinical usefulness of the nomogram by calculating the net benefits at different threshold probabilities (0%-100%). Statistical analyses were performed with SPSS software (Version 26.0, SPSS Chicago, USA), Stata software (V.15.0), and GraphPad Prism 8.0.

## Results

### Clinical characteristics and laboratory parameters

Comparison of participants’ demographic and clinical characteristics among the two groups is presented in Table 1. In total, 506 patients (70% male; mean age, 61±11 years) with STEMI who underwent PCI were included in the study. The SR group consisted of 69 patients (age, 60.89±10.98 years) and the No-SR group included 437 patients (age, 61.23±11.10 years). The prevalence of the SR was 13.6%. Assessment of risk factors revealed there were less smoking in the No-SR group than the SR group (60.6% vs. 73.9%, *P* < 0.05). In addition, it was also reported that the No-SR group were more frequently classified into the Killip class ≥ II on admission (12.6% vs. 8.7%, *P* < 0.05). Compared with the No-SR group, the LVEF was significantly higher in the SR group [54 (51, 59) vs. 52 (50, 56), *P* < 0.05]. However, there was no difference between the two groups about the history of MI, previous PCI, previous stroke, SBP, DBP, and heart rate (*P* > 0.05 for all comparisons). Table 1 also summarizes the laboratory parameters. The levels of leukocyte count, neutrophil count, NLR, and FBG were higher in the No-SR group than the SR group (*P* < 0.05, for all). However, compared with the SR group, the level of lymphocyte count was significantly lower in the No-SR group (*P* < 0.001). No significant difference was witnessed in total cholesterol, triglyceride, LDL-C, HDL-C, serum creatinine, and hemoglobin between two study groups (*P* > 0.05, for all comparisons).

**Table 1.**
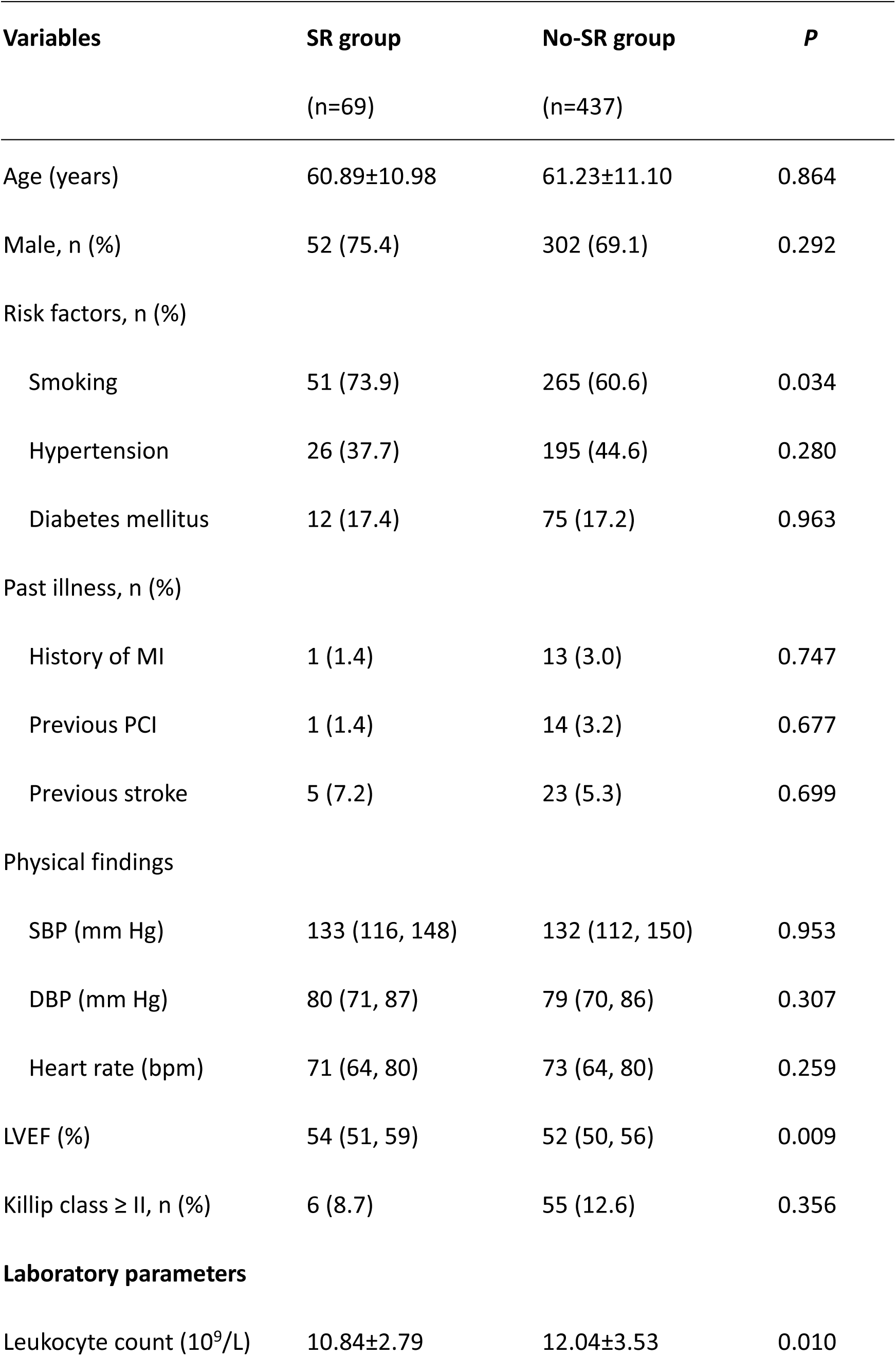
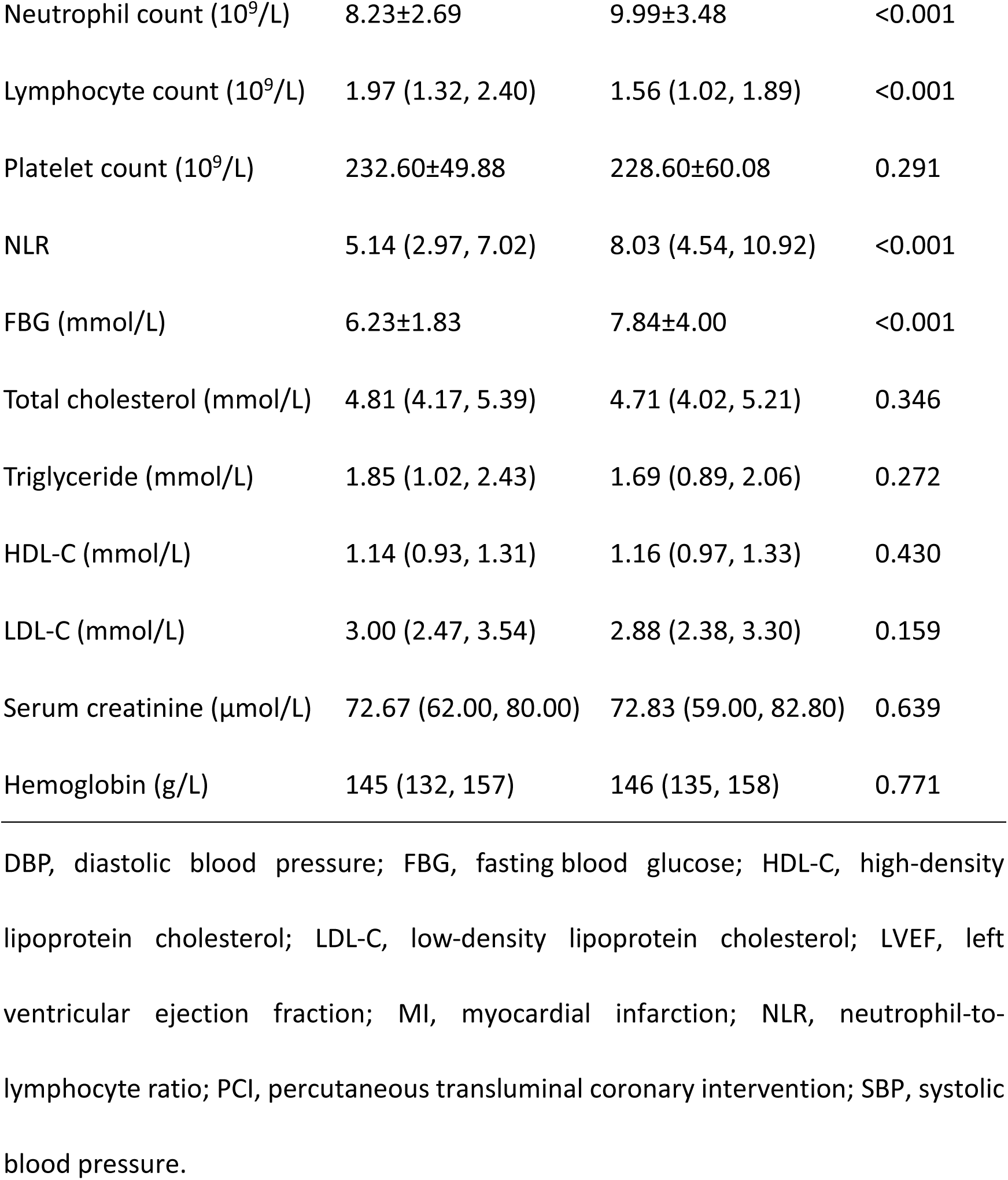
Basic characteristics of enrolling patients.

### Angiographic and treatment features

The SR group presented more proportions of final TIMI 3 flow than the No-SR group (98.6% vs. 91.5%, *P* < 0.05). There was no statistically significant difference in terms of infarct-related artery, myocardial wall, initial TIMI flow, stent length, stent diameter, stent number per patient, stenosed artery number, and multi-vessel disease between the two groups (*P* > 0.05 for all comparisons, Table 2).

**Table 2.** Angiographic, treatment, and procedural features

| **Variables** | **SR group** | **No-SR group** | **P** |
| --- | --- | --- | --- |
|  | (n=69) | (n=437) |  |
| Infarct-related artery, n (%) |  |  | 0.658 |
| LAD | 31 (44.9) | 177 (40.5) |  |
| LCX | 6 (8.7) | 52 (11.9) |  |
| RCA | 32 (46.4) | 208 (47.6) |  |
| Myocardial wall, n (%) |  |  | 0.897 |
| Anterior wall | 28 (40.6) | 175 (40.0) |  |
| Inferior wall | 39 (56.5) | 253 (57.9) |  |
| Others | 2 (2.9) | 9 (2.1) |  |
| Initial TIMI flow, n (%) |  |  | - |
| 0 | - | 379 (86.7) |  |
| 1 | - | 33 (7.6) |  |
| 2 | - | 25 (5.7) |  |
| 3 | 69 (100) | - |  |
| Final TIMI 3 flow, n (%) | 68 (98.6) | 400 (91.5) | 0.040 |
| Stenosed artery number, n (%) |  |  | 0.852 |
| 1 | 25 (36.2) | 154 (35.2) |  |
| 2 | 22 (31.9) | 154 (35.2) |  |
| 3 | 22 (31.9) | 129 (29.6) |  |
| Multi-vessel disease, n (%) | 44 (63.8) | 283 (64.8) | 0.873 |
| Thrombus aspiration, n (%) | 7 (10.1) | 207 (47.4) | <0.001 |
| Stent number per patient, n | 1 (1,1) | 1 (1,1) | 0.762 |
| Stent length (mm) | 28 (21, 33) | 28 (21, 35) | 0.546 |
| Stent diameter (mm) | 3.3 (2.7, 3.5) | 3.2 (2.7, 3.5) | 0.940 |
| Treatment n (%) |  |  |  |
| ACE-I/ARB | 44 (63.8) | 227 (51.9) | 0.067 |
| β- blocker | 40 (58.0) | 248 (56.8) | 0.849 |
| Spirolactone | 25 (36.2) | 150 (34.3) | 0.757 |

ACE-I, angiotensin-converting enzyme inhibitors; ARB, angiotensin receptor blockers; LAD, left anterior descending artery; LCX, left circumflex artery; RCA, right coronary artery; TIMI, thrombolysis in myocardial infarction.

### SR is associated with a favorable outcome

Assessment of complications during time of hospitalization revealed that 81 (18.5%) cases from the No-SR group developed congestive heart failure as compared with only 6 (8.7%) cases in the SR group (18.5% vs. 8.7%, *P* < 0.05). Moreover, the usage of mechanical ventilation was higher in the No-SR group than the SR group (*P* < 0.05). There was no significant difference between the two groups in the rate of in-hospital death (*P* > 0.05), however, rates of 6-month, 12-month, 18-month, 24-month and 30-month MACEs were higher in the No-SR group compared with the SR group (13.6% vs. 2.9%, *P* < 0.05;16.8% vs. 5.9%, *P* < 0.05; 19.9% vs. 7.2%, *P* < 0.05; 22.0% vs. 8.8%, *P* < 0.05; 23.4% vs. 10.3%, *P* < 0.05 respectively) (Table 3). Kaplan-Meier analysis revealed significantly poorer MACEs-free survival during follow-up in the No- SR group compared with the SR group (Log-rank test, *P* < 0.05), however, there was no statistical significance in death-free survival between the two groups (Log-rank test, *P* > 0.05) (Figure 1).

**Figure 1.**
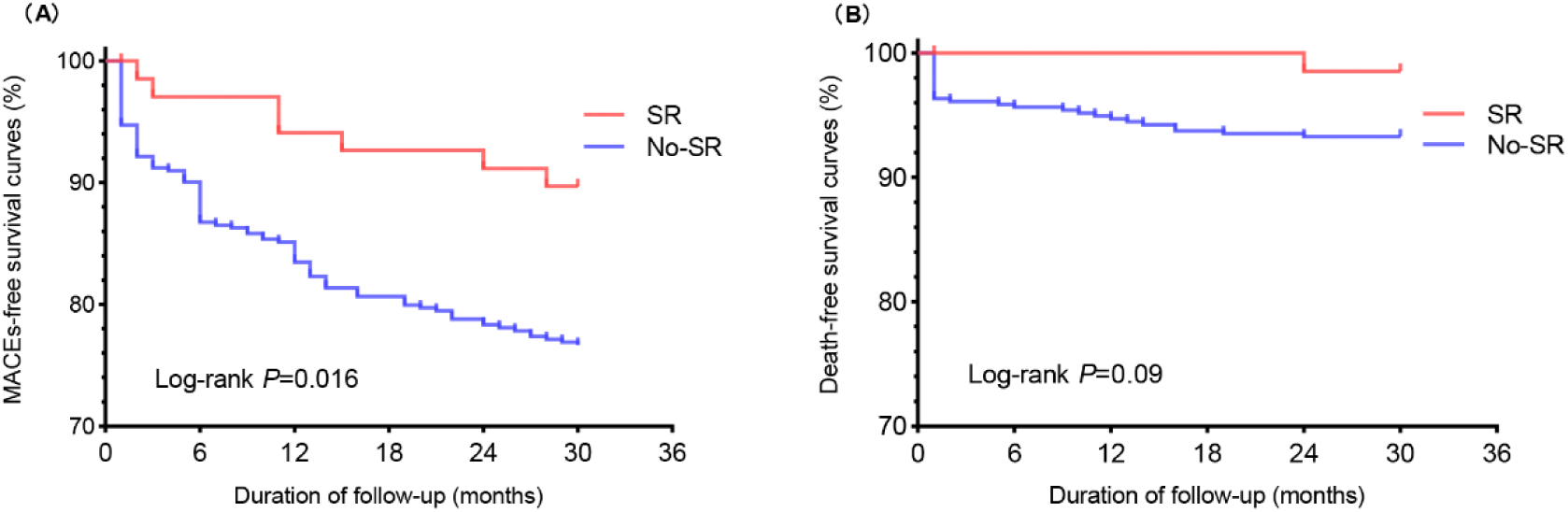
Kaplan-Meier survival curve analysis. (A) presents the MACEs-free survival curve and (B) reveals death-free survival curve during the follow-up. MACEs, major adverse cardiac events; No-SR, no spontaneous reperfusion (initial TIMI flow 0-2); SR, spontaneous reperfusion (initial TIMI flow 3).

**Table 3.** Clinical outcomes during in-hospital and follow-up

| **Variables** | **SR group** | **No-SR group** | **P** |
| --- | --- | --- | --- |
|  | (n=69) | (n=437) |  |
| IABP, n (%) | 0 (0) | 10 (2.3) | 0.371 |
| Mechanical ventilation, n (%) | 0 (0) | 36 (8.2) | 0.009 |
| Complication, n (%) |  |  |  |
| CHF | 6 (8.7) | 81 (18.5) | 0.041 |
| Arrhythmia | 9 (13.0) | 77 (17.6) | 0.347 |
| Length of stay (days) | 6 (5,7) | 6 (5,7) | 0.879 |
| In-hospital death, n (%) | 0 (0) | 15 (3.4) | 0.242 |
| 6-month follow-up, n (%) | n=68 | n=428 |  |
| All-cause death | 0 (0) | 19 (3.8) | 0.091 |
| MACEs | 2 (2.9) | 58 (13.6) | 0.013 |
| 12-month follow-up, n (%) | n=68 | n=428 |  |
| All-cause death | 0 (0) | 24 (4.8) | 0.060 |
| MACEs | 4 (5.9) | 72 (16.8) | 0.020 |
| 18-month follow-up, n (%) | n=68 | n=428 |  |
| All-cause death | 0 (0) | 27 (6.3) | 0.038 |
| MACEs | 5 (7.2) | 85 (19.9) | 0.012 |
| 24-month follow-up, n (%) | n=68 | n=428 |  |
| All-cause death | 1 (1.5) | 29 (6.8) | 0.103 |
| MACEs | 6 (8.8) | 94 (22.0) | 0.012 |
| 30-month follow-up, n (%) | n=68 | n=428 |  |
| All-cause death | 1 (1.5) | 29 (6.8) | 0.103 |
| MACEs | 7 (10.3) | 100 (23.4) | 0.015 |

CHF, congestive heart failure; IABP, intra-aortic balloon pump; MACEs, Major adverse cardiac events.

### NLR is an important indicator predicting poor TIMI flow (No-SR)

To identify predictors for SR, we further applied the logistic regression analysis. Variables such as smoking, LVEF, leukocyte count, neutrophil count, lymphocyte count, NLR, and FBG were entered into multivariate logistic regression analysis. Multivariate logistic regression analysis revealed NLR (OR: 0.799, 95% CI: 0.730-0.874, *P* < 0.001) and FBG (OR: 0.787, 95% CI: 0.683-0.906, *P* = 0.001) as the independent predictors of SR (Table 4).

**Table 4.** Predictors of SR flow by multivariate logistic regression analysis

| **Variables** | **B** | **Wald** | **OR** | **95% CI** | ***P*** |
| --- | --- | --- | --- | --- | --- |
| NLR | -0.224 | 23.918 | 0.799 | 0.730-0.874 | < 0.001 |
| FBG (mmol/L) | -0.240 | 11.061 | 0.787 | 0.683-0.906 | 0.001 |
| Smoking | 0.603 | 3.824 | 1.827 | 0.999-3.342 | 0.051 |

CI, confidence interval; FBG, fasting blood glucose; NLR, neutrophil-to-lymphocyte ratio; OR, odds ratio.

The association between NLR and initial TIMI flow were further corroborated by the analysis using P for trend and Per SD (Table 5). NLR was independently associated with No-SR flow after adjustment for variables in model 1. The result was preserved with the inclusion of variables in subsequent models 2, indicating that these variables did not act as mediators of the relationship between the NLR and No-SR. After multivariable adjustment ORs (95% CIs) across quartiles of NLR were 1.00 (reference), 1.261 (0.662, 2.404), 2.03 (1.002, 4.102), and 9.242 (3.081, 27.725) (P for trend, < 0.001); each SD of transformed NLR was associated with a 191% increment in OR of No-SR. More than that, we also employed ROC curve to study the predictive ability of NLR and FBG to poor TIMI flow (No-SR). The ROC curve showed that NLR had the higher area under the curve (AUC) (AUC = 0.685) than FBG (AUC = 0.629) (Figure 2). The optimal cut-off value of NLR was determined by Youden index from the ROC result. NLR at 7.22 was the best threshold with the optimal sensitivity (47.6%) and specificity (81.2%).

**Figure 2.**
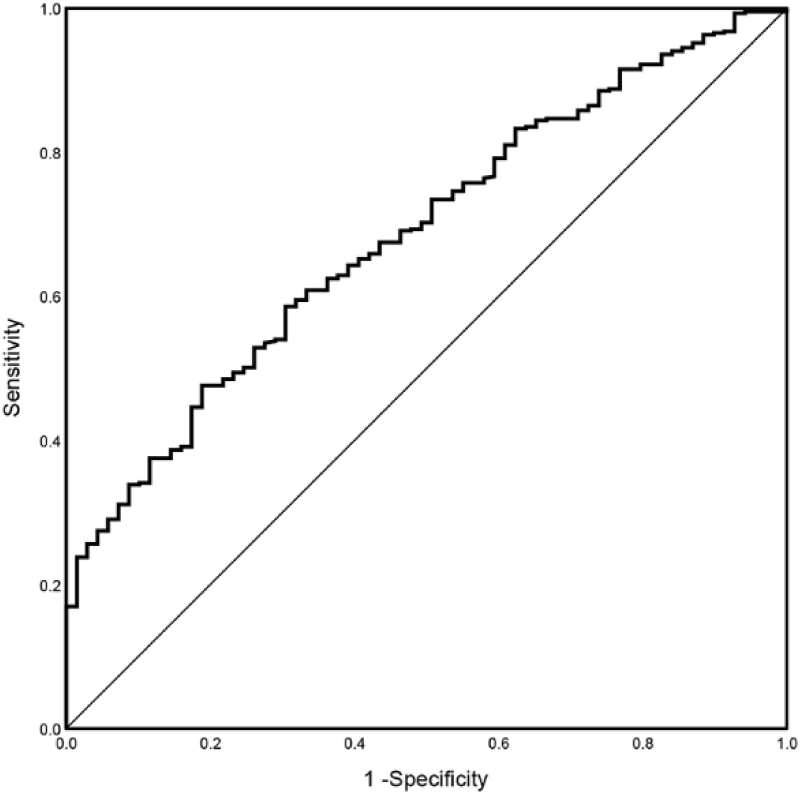
ROC curves of the diagnosis ability of NLR for No-SR. The optimal cut-off value of NLR for predicting No-SR was 7.22 with 47.6% sensitivity and 81.2% specificity. NLR, neutrophil-to-lymphocyte ratio; No-SR, no spontaneous reperfusion (initial TIMI flow 0-2); ROC, receiver operating characteristic.

**Table 5.** Association of NLR level with No-SR^1^

|  | Quartiles of NLR |  |  |  | *P* for trend | Per SD of transformed NLR | *P* |
| --- | --- | --- | --- | --- | --- | --- | --- |
|  | Q 1 | Q 2 | Q 3 | Q 4 |  |  |  |
| Variables | ≤4.23 | 4.24-6.67 | 6.58-10.17 | ≥10.18 |  |  |  |
| N (SR/ No-SR) | 28/99 | 22/104 | 15/111 | 4/134 |  |  |  |
| OR (95% CI) |  |  |  |  |  |  |  |
| Crude | 1 (Ref) | 1.337 (0.717-2.492) | 2.093 (1.057-4.114) | 8.697 (2.952-25.624) | *P*< 0.001 | 2.603 (1.736-3.902) | *P*< 0.001 |
| Model 1 | 1 (Ref) | 1.399 (0.746-2.624) | 2.259 (1.132-4.508) | 9.290 (3.138-27.503) | *P*< 0.001 | 3.049 (1.956-4.752) | *P*< 0.001 |
| Model 2 | 1 (Ref) | 1.261 (0.662-2.404) | 2.030 (1.002-4.102) | 9.242 (3.081-27.725) | *P*< 0.001 | 2.907 (1.086-7.783) | *P*< 0.05 |

<sup>1</sup> Model 1 was a logistic regression that was adjusted for age, sex, smoking, hypertension, diabetes mellitus, history of MI, previous PCI, previous stroke, SBP, DBP, heart rate, LVEF, and Killip class. Model 2 was a logistic regression that was adjusted as for model 1 and for leukocyte count, neutrophil count, lymphocyte count, platelet count, FBG, total cholesterol, triglyceride, HDL-C, LDL-C, serum creatinine, and hemoglobin.
CI, confidence interval; DBP, diastolic blood pressure; FBG, fasting blood glucose; HDL-C, high-density lipoprotein cholesterol; LDL-C, low-density lipoprotein cholesterol; LVEF, left ventricular ejection fraction; MI, myocardial infarction; NLR, neutrophil-to-lymphocyte ratio; OR, odds ratio; PCI, percutaneous transluminal coronary intervention; Q, quartile; Ref, reference; SBP, systolic blood pressure; SD, standard deviation.

### Construction and validation of nomogram model of predicting poor TIMI flow (No-SR)

The basic characteristics of different cohorts are described in Table S1. In order to get a more comprehensive view of the association between NLR and initial TIMI flow, we further performed a logistic regression analysis and constructed a predictive model. The results of the logistic regression analysis are showed in Table S2 and visualized in the form of a nomogram plot to help practice in the clinic (Figure 3). The AUC of this predictive model in the training cohort and validation cohort was 0.704 (95% CI: 0.633–0.775) and 0.747 (95% CI: 0.649–0.845) respectively (Figure S1), which suggested the good prediction capability of this model. The prediction accuracy of the nomogram was evaluated by the Hosmer–Lemeshow chi-square statistic calibration method and revealed that the *P* value in training cohort and validation cohort was 0.777 and 0.334 respectively (Figure S2), which confirms that the model fits nicely. Figure S3 describes the DCA of the prediction nomogram, which reveals that the nomogram can achieve good net benefit.

**Figure 3.**
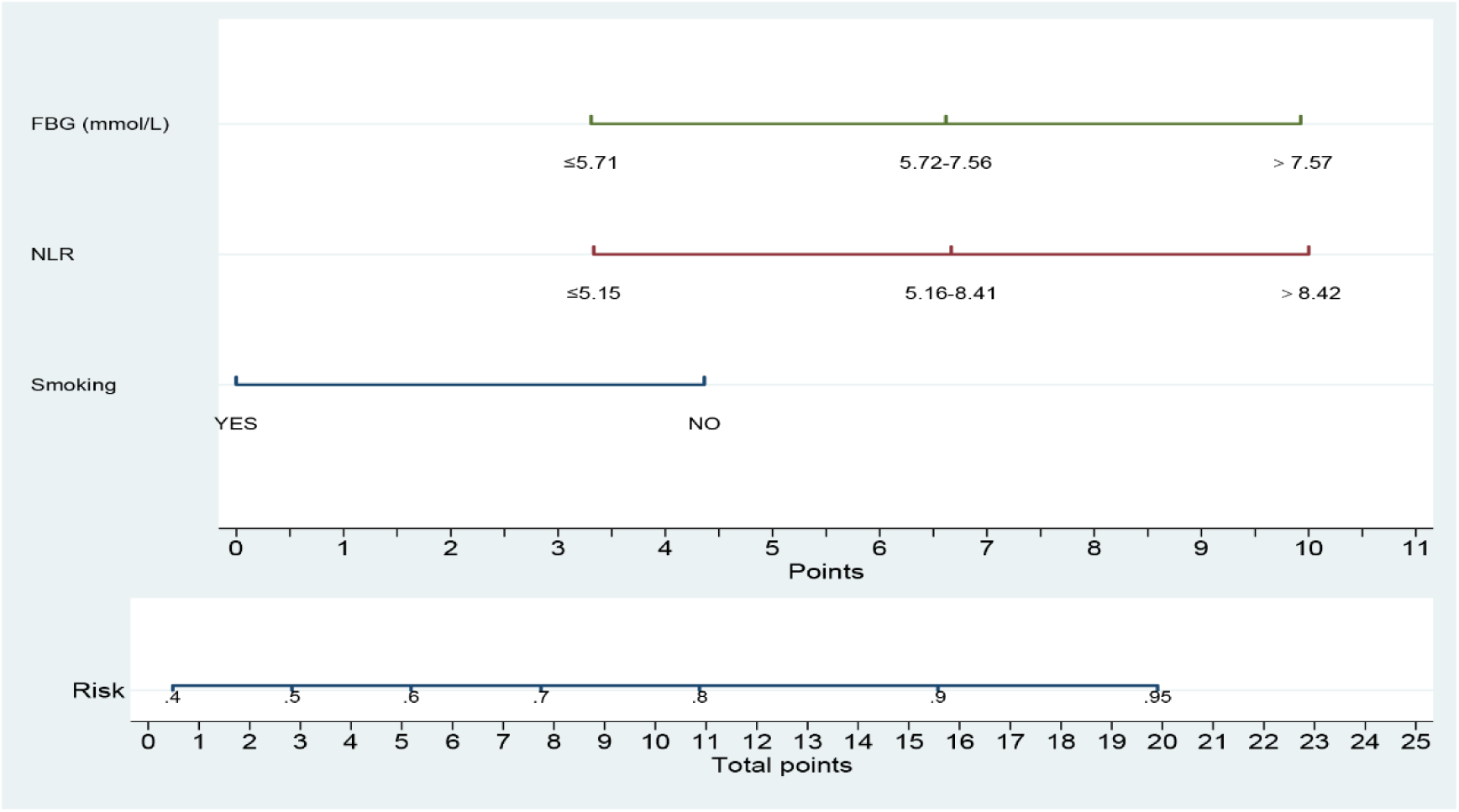
Nomogram for predicting No-SR. The present nomogram indicates the possible risk prediction of coronary initial No-SR. First to third rows: predictors of No- SR; fourth row: point assignment of the variables; fifth row: prediction of the risk of No-SR; sixth row: total score of three predictors. FBG, fasting blood glucose; NLR, neutrophil-to-lymphocyte ratio; No-SR, no spontaneous reperfusion (initial TIMI flow 0-2).

### NLR is an important indicator predicting MACEs during follow-up

Median follow-up duration was 30 months, corresponding to 1055 person-years. The MACE rate was 10% person- years. In this study, we further analyzed the hazard ratio of indicators in MACEs during follow-up in the overall population by univariate Cox regression (Table 6), and the multivariate Cox regression analysis revealed NLR (HR: 1.035, 95% CI: 1.001-1.071, *P* < 0.05) rather than SR was the independent predictor (Figure 4). To further determine the association of NLR and MACEs, all patients were divided into low NLR group (NLR ≤ 6.57, n = 253) and high NLR group (NLR > 6.57, n = 253) according to the median of NLR. Kaplan-Meier analysis shows significantly poorer MACEs-free survival in the high NLR group compared with the NLR low group (Log-rank test, *P* < 0.05) (Figure 5).

**Figure 4.**
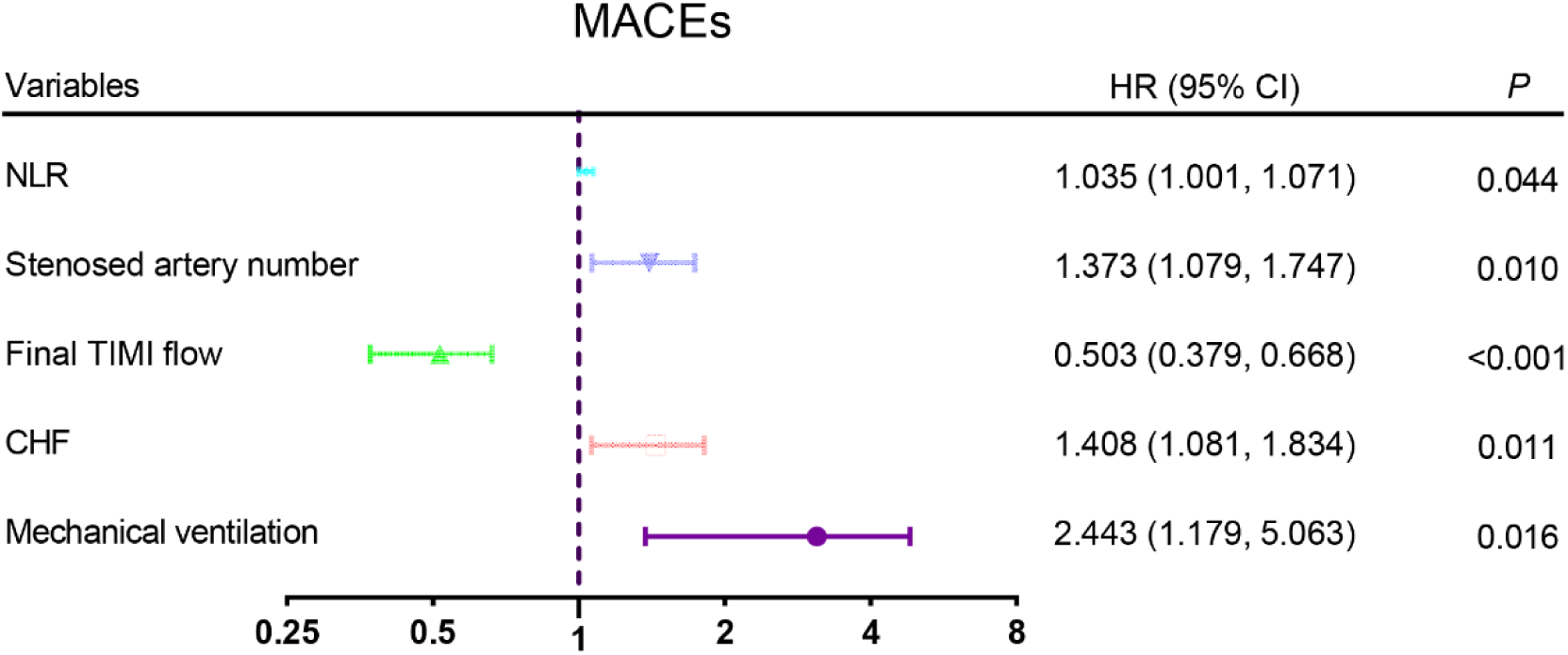
Independent risk factors of MACEs during follow-up by multivariate Cox regression analysis. It shows that NLR, stenosed artery number, final TIMI flow, CHF, and mechanical ventilation are independent risk factors of MACEs during follow-up in STEMI patients. CHF, congestive heart failure; HR, hazard ratio; MACEs, major adverse cardiac events; NLR, neutrophil-to-lymphocyte ratio; STEMI, ST-segment elevation myocardial infarction.

**Figure 5.**
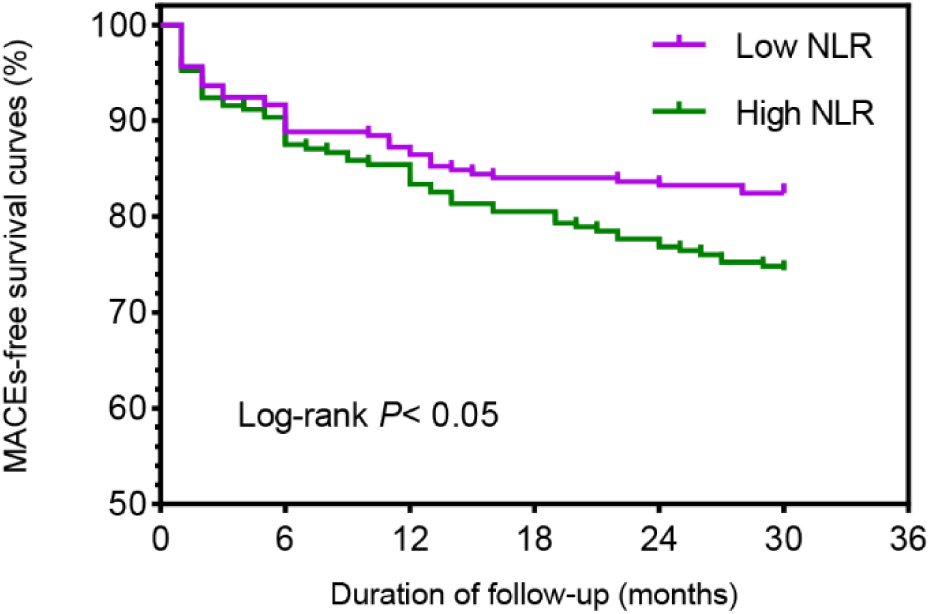
Kaplan-Meier survival curve analysis. MACEs-free survival (Log-rank test, P < 0.05) according NLR distribution divided by medina. Transformed NLR are the following: Low NLR: ≤ 6.57; High NLR > 6.57. MACEs, major adverse cardiac events; NLR, neutrophil-to-lymphocyte ratio.

**Table 6.**
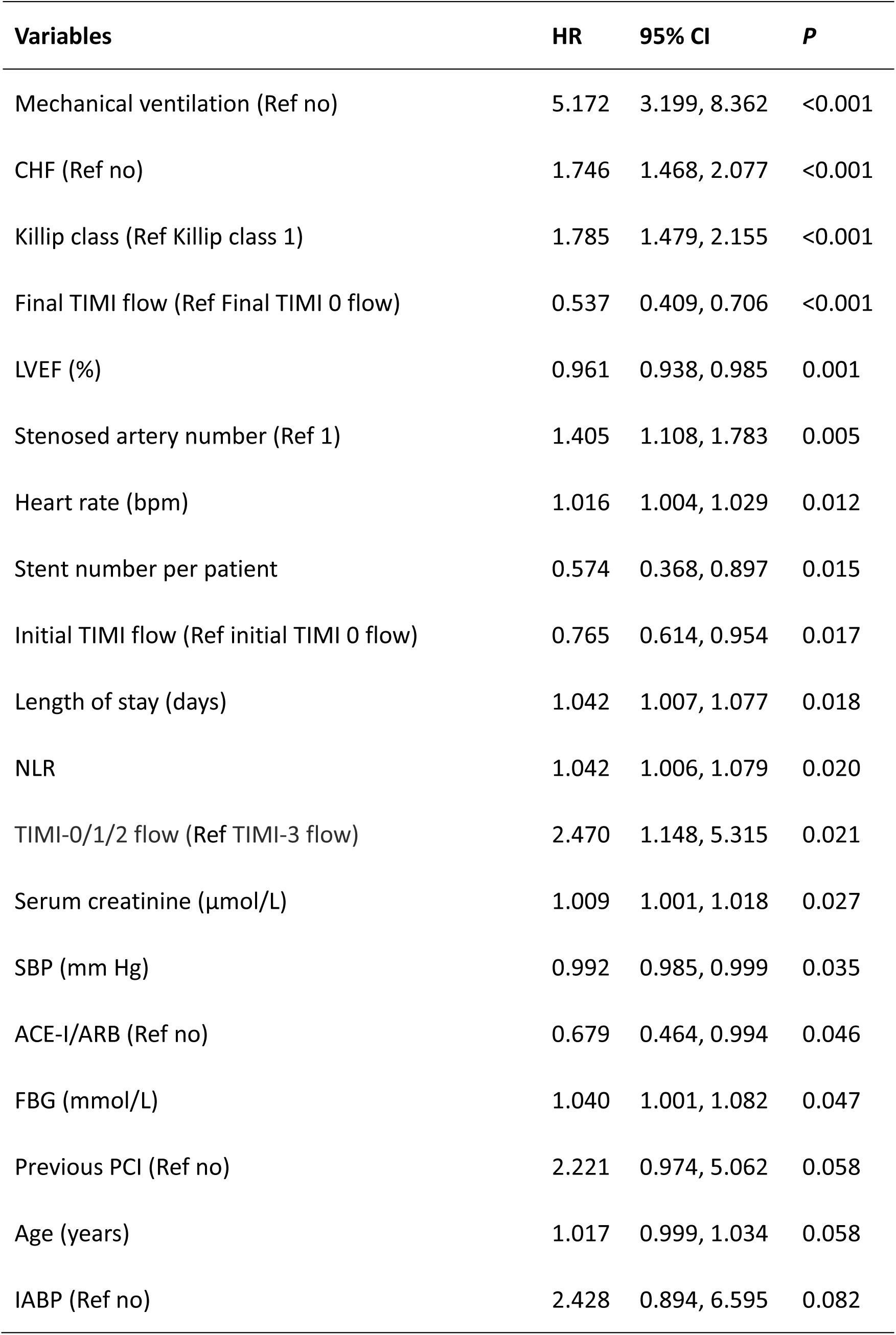
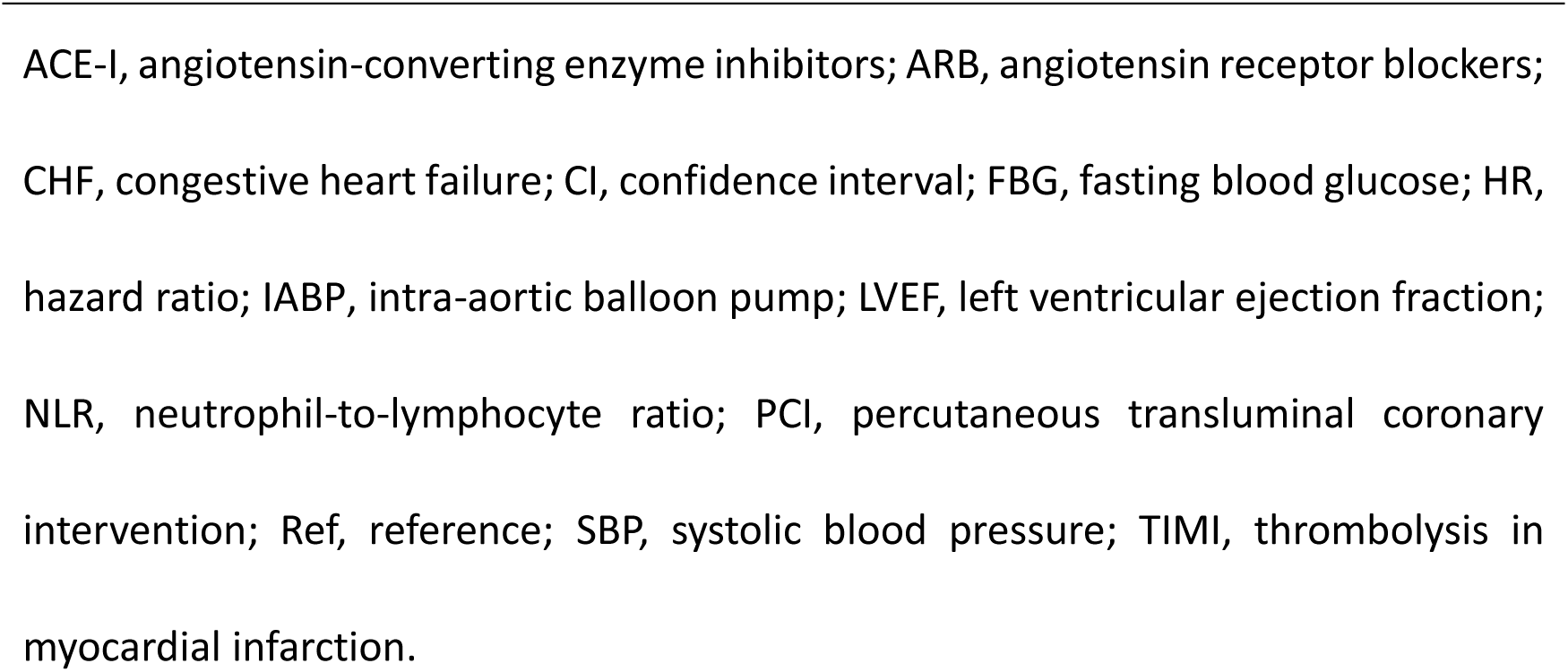
Univariable Cox regression analysis for predicting the MACEs

| **Variables** | **HR** | **95% CI** | **P** |
| --- | --- | --- | --- |
| Mechanical ventilation (Ref no) | 5.172 | 3.199, 8.362 | <0.001 |
| CHF (Ref no) | 1.746 | 1.468, 2.077 | <0.001 |
| Killip class (Ref Killip class 1) | 1.785 | 1.479, 2.155 | <0.001 |
| Final TIMI flow (Ref Final TIMI 0 flow) | 0.537 | 0.409, 0.706 | <0.001 |
| LVEF (%) | 0.961 | 0.938, 0.985 | 0.001 |
| Stenosed artery number (Ref 1) | 1.405 | 1.108, 1.783 | 0.005 |
| Heart rate (bpm) | 1.016 | 1.004, 1.029 | 0.012 |
| Stent number per patient | 0.574 | 0.368, 0.897 | 0.015 |
| Initial TIMI flow (Ref initial TIMI 0 flow) | 0.765 | 0.614, 0.954 | 0.017 |
| Length of stay (days) | 1.042 | 1.007, 1.077 | 0.018 |
| NLR | 1.042 | 1.006, 1.079 | 0.020 |
| TIMI-0/1/2 flow (Ref TIMI-3 flow) | 2.470 | 1.148, 5.315 | 0.021 |
| Serum creatinine ($\mu\text{mol/L}$) | 1.009 | 1.001, 1.018 | 0.027 |
| SBP (mm Hg) | 0.992 | 0.985, 0.999 | 0.035 |
| ACE-I/ARB (Ref no) | 0.679 | 0.464, 0.994 | 0.046 |
| FBG (mmol/L) | 1.040 | 1.001, 1.082 | 0.047 |
| Previous PCI (Ref no) | 2.221 | 0.974, 5.062 | 0.058 |
| Age (years) | 1.017 | 0.999, 1.034 | 0.058 |
| IABP (Ref no) | 2.428 | 0.894, 6.595 | 0.082 |

ACE-I, angiotensin-converting enzyme inhibitors; ARB, angiotensin receptor blockers; CHF, congestive heart failure; CI, confidence interval; FBG, fasting blood glucose; HR, hazard ratio; IABP, intra-aortic balloon pump; LVEF, left ventricular ejection fraction; NLR, neutrophil-to-lymphocyte ratio; PCI, percutaneous transluminal coronary intervention; Ref, reference; SBP, systolic blood pressure; TIMI, thrombolysis in myocardial infarction.

## Discussion

The current study not only evaluated the relationship between NLR and SR, but also demonstrated the association of NLR and long-term prognosis.

The main conclusions of this analysis are the following: first, NLR level is an important indicator predicting occurrence of SR in STEMI patients, which is associated with a favorable outcome; second, significantly poor long-term prognosis in higher NLR patients compared with the lower NLR patients although they all received timely percutaneous coronary intervention; third, NLR an independent indicator predicting long-term prognosis.

Early achievement of SR in IRA plays a major role to obtain favorable prognosis after STEMI. ^6^ Patients presenting initial TIMI-3 flow have better clinical outcomes (e.g., limiting the size of infarction, low incidence of heart failure, lower leukocyte infiltration, high proportions of final TIMI 3 flow) after PCI than those without it. ^6,17^ Thus, at the early stage of STEMI, the patency of IRA influences the clinical course after reperfusion. Overall, early detection of SR might be crucial for clinical decision making after STEMI.

The incidence of SR was 13.6% in this study, which is consistent with previous reports. ^18^ This study result is consistent with other published studies that the SR patients with STEMI have a higher proportion of final 3 flow, better prognosis, and a lower incidence of heart failure, thrombus burden, and lower levels of leukocyte than the No-SR group. ^6,19,20^ However, few studies have explored the relationship between inflammation biomarkers (NLR) and SR, prognosis. In order to demonstrate the correlation between NLR and SR, we applied univariate analysis, P for trend and Per SD analysis, finally we drew a similar conclusion, i.e., NLR has a negative correlation with SR, what’s more, NLR is an independent predictor for SR.

Previous studies have shown that acute myocardial infarction can lead to a strong inflammatory response, i.e., abundant leukocytes infiltrate into the infarcted heart and adhere to cardiomyocytes and exert cytotoxic effects, exacerbating ischemic myocardial injure. ^21^ The process of the close spatial interaction between leukocytes and viable cardiomyocytes and the consequent cardiomyocyte injury gave rise to the notion of leukocyte-mediated cardiomyocyte injury. Neutrophil as the most important subgroup of leucocyte, recently is regarded as an emerging biomarker in cardiovascular diseases, which is characterized by mediating the progression of atherosclerotic plaque and balancing tissue damage. It is a known fact that the rupture of the vulnerable atherosclerotic plaque leading to occlusion of the coronary artery is the primary mechanism of STEMI. During the process of plaque rupture, neutrophils aggregate in the vessel wall, and infiltrate into the atherosclerotic plaques mediating destabilization of atherosclerotic plaques (e.g., decreasing collagen content, a marker of stable plaque, thus resulting in rupture of plaque). ^9,10,22^ In addition, neutrophils can also interact with endothelial cells, roll along the vascular wall, become activated, pass through the vessel wall, infiltrate the infarcted area, adhere to surviving cardiomyocytes, and ultimately produce reactive oxygen species, resulting in severe cytotoxic effects and aggravating cardiomyocyte injury. ^21,23,24^ These processes culminate in the damages of cardiac structure and function. Lymphocytes have been reported to play a significant role in mediating the inflammatory response at all phases of the atherosclerotic process and can lead to rupture of atherosclerotic plaques. ^9^ The lymphopenia of AMI patients reflects the excessive apoptosis of lymphocytes in the state of inflammation ^25^ and the dysfunction of immune regulation. ^26^ In addition, the relative lymphopenia is also mediated by the increased endogenous cortisol.^25^

Both neutrophil and lymphocyte have been proven to be associated with cardiovascular diseases, however, NLR, as an integration of neutrophil and lymphocyte, has been demonstrated superior to neutrophil or lymphocyte alone. Recently, increased NLR was reported to be associated with short-term prognosis of AMI, ^14^ malignant tumors, ^27^ stroke, ^13^ infectious diseases, ^28^ and other diseases. As neutrophils and lymphocytes play an important role in the immune response of acute myocardial infarction, thus, NLR becomes an independent factor in accurately predicting SR and contributes to the risk stratification of patients with STEMI. We established a model for predicting SR, which included three critical indicators, namely, NLR, glucose and smoking. These three indicators are easy, convenient and economical to obtain, which makes it easier to identify high-risk STEMI patients and has definite significance in guiding clinical assessment.

Our study found that the proportion of thrombus aspiration in No-SR patients was significantly increased, indicating that the thrombus burden in No-SR patients was overweight. Rupture of the vulnerable atherosclerotic plaque leading to occlusion of the coronary artery could set off an uncontrolled cascade of thrombus formation and leukocyte infiltration, which in turn, aggravate the formation of thrombus. That would one explanation why the level of inflammatory markers in SR patients were lower. In addition, with increased coronary thrombus burden, STEMI patients have a relatively high no-reflow risk. ^29^

Final 0-2 flow known as no-reflow phenomenon, ^30^ STEMI patients with slow-flow/no-reflow have adverse prognosis than patients with normal flow. ^31^ Consistent with reports in other study, ^6^ our research demonstrated that the proportion of final 3 flow in SR patients was higher than that of the No-SR patients. Possibly it reflects a negative correlation between spontaneous reperfusion (SR) and no-reflow phenomenon (less final 3 flow). The underlying mechanisms of no-reflow phenomenon are the dysfunction and structural disruption of coronary microcirculation, including the leukocyte aggregation, microembolization, microvascular vasomotion, and cellular oedema. ^30^ Several studies have shown a relationship between no-reflow phenomenon and larger infarct size. ^30,32,33^ No-reflow phenomenon is linked to prolonged ischemia, further leading to larger infarct size, lower left ventricular ejection fraction, and increased incidences of heart failure, and death. ^30^ In order to establish the relationship between prognosis and no-reflow phenomenon, we applied Cox regression analysis and found final TIMI flow was an independent indicator predicting MACEs during follow-up. Similar studies also showed that 5-year mortality was twice as high in patients with STEMI with poor final TIMI flow compared with patients with normal flow. ^31^

The present study had demonstrated the No-SR patients were more frequently classified into the Killip class ≥ II on admission, suffering from higher prevalence of heart failure with lower LVEF, which was similar to previous studies. ^6,34^ When the initial TIMI flow in the STEMI patients is abnormal, the heart suffered prolonged ischemia, causing irreversible cardiomyocyte and endothelial damage, microcirculation dysfunction, leukocyte infiltration. ^30^ The damage, in turn, leads to severe ischemic insult and forms a vicious circle, eventually resulting in severe myocardial ischemic injury and heart failure. As mentioned above, No-SR patients were more likely to a higher proportion of no-reflow phenomenon, which might another reason No-SR was associated with heart failure. Patients who develop heart failure in the setting of AMI bear adverse prognosis, ^35-37^ which is about four times more likely to die in hospital compared with those who without heart failure. ^37^ This could explain why heart failure was an independent predictor of MACEs during follow-up as we demonstrated in our study.

Inflammation not only plays an important role in plaque rupture in acute myocardial infarction, but also influences myocardial remodeling and prognosis after myocardial infarction (Figure 6). Potential mechanisms could be as follows: First, significantly increased NLR aggravates the occurrence of No-SR and no-reflow phenomenon, while elevated No-SR also increases the risk of no-reflow phenomenon, which has the potential to expand the infarction size and leads to heart failure, resulting in poor prognosis. Second, in addition, elevated No-SR and NLR can also directly aggravate myocardial injury and culminate in poor prognosis. Furthermore, not only NLR, but also heart failure and final TIMI flow are also independent risk predictors for MACEs during follow-up. NLR can also be used as an indicator for long-term prognosis.

**Figure 6.**
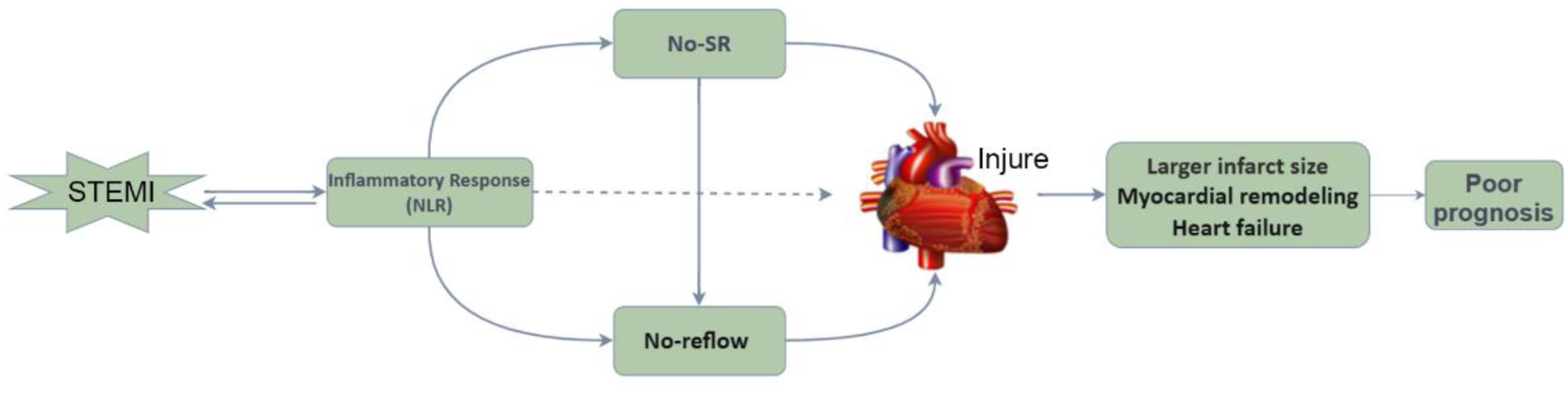
Association of inflammatory injury and myocardial infarction. Acute myocardial infarction can lead to a strong inflammatory response. Inflammation plays a major role in plaque instability, rupture, and erosion, and facilitates its progression, leading to thrombus formation, and resulting in coronary artery occlusion and myocardial infarction. Neutrophils interact with endothelial cells, roll along and pass through the vessel wall, infiltrate the infarcted area, adhere to surviving cardiomyocytes, and ultimately produce reactive oxygen species, resulting in severe cytotoxic effects and aggravating cardiomyocyte injury. Leukocyte aggregation is also one of the underlying mechanisms of no-reflow phenomenon. Elevated No-SR also increased the risk of no-reflow phenomenon. Inflammatory injury, no-reflow phenomenon, and No-SR together culminate in the damages of cardiac structure and function, leading to larger infarct size, lower left ventricular ejection fraction, and increased incidences of heart failure, and death. NLR, neutrophil-to-lymphocyte ratio; No-SR: no spontaneous reperfusion (initial TIMI flow 0-2); STEMI, ST-segment elevation myocardial infarction.

Some researches had shown the association between NLR and AMI. A small sample study reported post-PCI NLR could predict adverse prognosis in AMI, ^38^ however, the study did not distinguish between STEMI and NSTEMI. As we known, patients with NSTEMI usually have a favorable prognosis than STEMI individuals. Another relatively large clinical study analyzed the role of NLR in predicting the short-term prognosis of NSTEMI and STEMI respectively ^14^, the study included all AMI patients, regardless of whether they received PCI treatment. Unlike previous researches, our study explored the role of NLR in predicting long-term prognosis in STEMI patients who received primary PCI.

NLR can identify high-risk patients with myocardial infarction, and become an independent factor influencing the poor prognosis of myocardial infarction, although all the patients received primary percutaneous coronary intervention, indicating that NLR plays an important role in acute myocardial infarction. The attribute of NLR (e.g., easy, convenient, and economical to obtain) also determine that it will be widely used in clinic.

## Limitations

The present study has several limitations. First, this is a single-centered with retrospective design and our observational study was based on a single center which may cause selection bias. Second, we did not collect other inflammation biomarkers, such as IL-1 value, IL-6 value, TNF-a value, and CRP value. Third, when we constructed the nomogram model for predicting poor TIMI flow, we only included internal validation and external validation was lacked.

## Conclusions

In conclusion, NLR had the ability in predicting SR in STEMI patients and SR was associated with a favorable outcome. We also revealed an association between NLR and increased risk of MACEs during follow-up. Therefore, this study provided new evidence for the role of NLR in the risk classification of STEMI patients.

## Supplemental Material

Tables S1–S2

Figures S1-S3

## Data Availability

Data available on request from the authors.

## Acknowledgement

We thank the personnel at our centers participating in the research for their commitment and professionalism to this study.

## Author Contributions

Bing was responsible for writing the manuscript; Collection, analysis, or interpretation of data: Bing Li, Mingyou Zhang and Yaoting Zhang; Yang Zheng and He Cai were responsible for supervision. All authors read and approved the final manuscript.

## Nonstandard Abbreviations and Acronyms

FBG: fasting blood glucose
IRA: infarct-related artery
LVEF: left ventricular ejection fraction
MACEs: Major adverse cardiac events
NLR: neutrophil-to-lymphocyte ratio
No-SR: no spontaneous reperfusion (initial TIMI flow 0-2)
SR: spontaneous reperfusion (initial TIMI flow 0-3)

